# Facial Emotion Processing and GABAergic Modulation in Individuals at Clinical High-Risk for Psychosis

**DOI:** 10.64898/2026.11.12.26367443

**Authors:** Nikita C Julius, Paulina B Lukow, Amanda Kiemes, Nicholas R Livingston, Maria-Mihaela Avram, Samuel R Knight, Peter CT Hawkins, Owen O’Daly, Luke Jelen, Thomas J Reilly, Aikaterini Dima, Maria A Nettis, Cecilia Casetta, Natasha Vorontsova, James Davies, Matthew B Wall, Thomas J Spencer, Andrea de Micheli, Paolo Fusar-Poli, Anthony A Grace, Kosha Ruparel, Ruben C Gur, Fernando Zelaya, Steven CR Williams, Philip McGuire, Federico Turkheimer, Mitul A Mehta, Edward Chesney, Gemma Modinos

## Abstract

Deficits in social cognition, including facial emotion recognition (FER), are present in individuals at clinical high-risk for psychosis (CHR-P). Altered GABAergic and glutamatergic signalling within corticolimbic circuitry has been proposed to underlie FER deficits in psychosis. We investigated FER performance and brain activation in CHR-P individuals, and whether pharmacological enhancement of GABAergic signalling with diazepam modulates these processes. In Study 1, 24 antipsychotic-naïve CHR-P individuals and 24 healthy controls underwent fMRI while performing the Penn Emotion Recognition Task. In Study 2, 24 antipsychotic-naïve CHR-P individuals participated in a randomised, double-blind, placebo-controlled, crossover study of 5mg diazepam versus placebo. Threat-related FER performance and neural activation were compared between groups and drug conditions. In Study 1, CHR-P individuals showed longer reaction times across emotions (n=22 vs n=23, *p*=0.01) and greater amygdala (*p*_FWE_=0.01) and hippocampal (*p*_FWE_=0.04) activation for neutral>fixation. Lower amygdala activation for threat>neutral (*p*_FWE_=0.03) and non-threat>neutral (*p*_FWE_=0.01), and lower hippocampal activation for non-threat>neutral (*p*_FWE_=0.04) may reflect this elevated response to neutral faces. Follow-up analyses showed lower insula (*p*_FWE_=0.01) and medial prefrontal cortex activation (*p*_FWE_=0.02) for threat>fixation. In Study 2, no significant drug effect on sensitivity was observed in the full sample (n=23), although secondary reduced-sample analyses, excluding one participant identified as influential, suggested improved overall sensitivity under diazepam (n=22; *p*=0.0499). After adjusting for global cerebral blood flow, diazepam reduced midcingulate cortex activation during neutral>fixation (pFWE=0.04). These findings highlight altered neural processing of neutral expressions in individuals at CHR-P, and suggest that aspects of emotion processing may be sensitive to GABAergic modulation.

## 1. Introduction

Deficits in social cognition, particularly in interpreting socio-emotional cues, are a well-established feature of schizophrenia-spectrum disorders (SSDs) [1–4]. Difficulties with facial emotion recognition (FER) are associated with poor social and functional outcomes [5, 6], and may also contribute to the pathophysiology of psychosis. Aberrant processing of emotionally salient social cues, particularly threat-related, has been implicated in psychotic symptoms such as persecutory delusions [7], which are reported in about 55-77% of children and adolescents at clinical high-risk for psychosis (CHR-P) presenting to mental health services [8, 9]. Thus, emotion processing may be an important domain for understanding both the clinical and functional manifestations of psychosis.

The CHR-P state describes individuals at elevated risk of developing psychosis, approximately 25% of whom transition to psychosis within 3 years of clinical presentation [10]. Individuals at CHR-P show moderate social-cognitive deficits relative to healthy individuals [4], including difficulties in recognising neutral and threat-related facial expressions such as anger, disgust, and fear [4, 11, 12]. A recent meta-analysis further found more pronounced baseline emotion processing deficits in CHR-P individuals who subsequently transitioned to psychosis than those who did not [11], suggesting that characterising FER deficits and their neural correlates may provide insights into mechanisms associated with psychosis risk (14-18).

Functional neuroimaging studies in SSDs have implicated altered recruitment of corticolimbic regions during FER, including the amygdala, anterior cingulate cortex (ACC), fusiform gyrus, hippocampus, insula, and medial prefrontal cortex (mPFC) [13–16]. Findings in CHR-P have been less consistent. A recent meta-analysis identified convergent alterations only in the posterior cingulate cortex at a lenient threshold [17], while individual studies have reported alterations in prefrontal, insular, thalamic, and inferior frontal regions [18–21]. These findings raise the possibility of altered corticolimbic recruitment during FER in CHR-P, although the underlying neurobiological mechanisms remain unclear.

One proposed mechanism is disrupted balance between excitatory glutamatergic and inhibitory gamma-aminobutyric acid (GABA)-ergic neurotransmission [22]. In the hippocampus, reduced GABAergic interneuron function has been proposed to contribute to excitatory/inhibitory imbalance in SSDs [23–27]. Given its reciprocal connectivity with other corticolimbic regions involved in emotion processing, including the amygdala [28–31], altered hippocampal GABAergic function may affect broader network function [13, 17, 32]. GABAergic signalling can be pharmacologically modulated with benzodiazepines such as diazepam, a positive allosteric modulator at the GABA_A_ receptor that potentiates the effects of endogenous GABA [33]. Benzodiazepines have been shown to modulate FER in healthy individuals [34–36], supporting a role for GABAergic inhibition in the neural mechanisms underlying emotion processing. Enhancing GABAergic signalling may modulate FER performance and its neural circuitry, providing a pharmacological probe of GABAergic contributions to emotion-processing abnormalities in CHR-P.

Accordingly, we sought to determine whether threat-related FER is altered in individuals at CHR-P and whether it is modifiable through acute GABAergic modulation. Signal Detection Theory (SDT) analyses were used to distinguish sensitivity from response bias [37, 38]. In Study 1, we examined behavioural performance and corticolimbic activation during threat-related FER in antipsychotic-naïve individuals at CHR-P and healthy controls (HCs). We hypothesised that CHR-P individuals would show heightened threat processing and aberrant corticolimbic activation [7, 11, 39–42]. In Study 2, we investigated whether 5 mg diazepam modulated threat-related FER in CHR-P. We hypothesised that diazepam would attenuate threat-related processing at behavioural and neural levels.

## 2. Methods

### 2.1 Participants: Study 1

Twenty-four antipsychotic-naïve individuals at CHR-P aged 18-40 years were recruited from OASIS (Outreach and Support in South London), an early-intervention service within the South London and Maudsley NHS Foundation Trust, UK [43]. CHR-P status was confirmed using the Comprehensive Assessment of At-Risk Mental States (CAARMS) [44] and participants were required to be currently experiencing attenuated psychosis symptoms, defined as scoring ≥3 on the severity and frequency scoring system on P1-P4 of the CAARMS. Twenty-four HCs aged 18-40 years were recruited through public advertisements within the same geographical region. All participants were required to have capacity to consent, IQ>70 (Wechsler Adult Intelligence Scale III, shortened version [45]), no history of neurological disorders, and no current or past exposure to antipsychotic medication. Additionally, HCs were required to have no lifetime history of an Axis-I psychiatric disorder or first-degree relative with such a disorder. Further eligibility criteria have been described elsewhere [46]. Thirteen participants took part in both Study 1 and Study 2; nine of these participants took part in Study 1 first. Participant sample sizes and exclusions are detailed in Supplementary Table S1.

### 2.2. Participants: Study 2

Twenty-four antipsychotic-naïve individuals at CHR-P aged 18-40 years were recruited from the OASIS to participate in a randomised, double-blind, placebo-controlled, crossover design study. CHR-P and eligibility criteria were as described for Study 1. Further eligibility criteria are described elsewhere [47]. Thirteen participants took part in both Study 1 and Study 2; four of these participants took part in Study 2 first. Participant sample sizes and exclusions are detailed in Supplementary Table S1.

### 2.3 Procedures

Both studies were conducted at King’s College London and received approval from the UK National Health Service Research Ethics Committees (Study 1: 17/LO/1130; Study 2: 18/LO/0618). Study 2 was registered on clinicaltrials.gov (NCT06190483). All participants provided written informed consent.

In Study 1, participants attended a screening visit followed by a single PET-MR scanning session during which they underwent fMRI while completing the Penn Emotion Recognition Task (ER40) [42, 48, 49]. Only ER40 fMRI data are analysed in the present study, further study procedure details can be found in the Supplementary Methods.

In Study 2, participants attended a screening visit and underwent two MRI sessions following a single oral dose of 5 mg diazepam or placebo (50 mg ascorbic acid) in a randomised, counterbalanced order, separated by a minimum 3-week washout period. Diazepam or placebo was administered 60 minutes before scanning, and participants completed the ER40 during blood oxygen-level dependent (BOLD) fMRI. Further procedural details are provided in the Supplementary Methods.

### 2.4 Emotion recognition task

The ER40 assesses recognition of static facial expressions [42, 48, 49]. Participants viewed 60 faces expressing anger, fear, happiness, sadness or neutral expressions for 4000 ms and identified the displayed emotion by button-press on a joystick. Variable intervals ranging from 2000-12000ms, weighted towards shorter intervals, displaying a complex fixation cross matched on facial perceptual qualities were interspersed throughout the trials. For analysis, anger + fear were combined as threat, happy + sad were combined as non-threat, and neutral expressions were modelled separately [48]. Full task details are provided in the Supplementary Methods.

### 2.5 MRI acquisition

MRI data were acquired at Perceptive Discovery, London, using a 3T GE SignaTM PET-MR General Electric scanner for Study 1 and at the Centre for Neuroimaging Sciences, King’s College London, using a General Electric MR750 3.0 T MR scanner with a 32-channel head coil for Study 2. T1-weighted structural and BOLD fMRI data were acquired in both studies. Full acquisition parameters for each study are provided in the Supplementary Methods.

We previously demonstrated that 5 mg diazepam reduced global cerebral blood flow (CBF) in this Study 2 sample [47]. Because drug-induced changes in global CBF may influence estimates of task-related BOLD activation [50], Study 2 analyses were repeated including global CBF as a covariate. CBF was quantified using pseudo-continuous arterial spin labelling as described previously [51–53], with further details provided in the Supplementary Methods.

### 2.6 Image preprocessing

#### 2.6.1 ER40 fMRI

Quality control of raw images was conducted using MRIQC [54]. Functional images were preprocessed using fMRIPrep 23.1 [55], including motion and slice timing correction, co-registration, and normalisation to the MNI template. Volumes with framewise displacement (FD)>1mm were modelled as regressors of no interest in first-level general linear models (GLM), and individual functional images were spatially smoothed with a 6mm FWHM Gaussian kernel using SPM12 [56]. Additional QC procedures [50] resulted in contrast-specific exclusions, but no complete participant exclusions. Further details are provided in the Supplementary Methods and Supplementary Table S1.

#### 2.6.2 ASL preprocessing: Study 2

Quantitative CBF maps for each participant were co-registered, segmented, and normalised to the MNI template and spatially smoothed (6mm FWHM) using the Automated Software for ASL Processing toolbox [57]. Visual quality control of individual CBF maps was conducted using MRIcroGL [58]. Mean global grey matter CBF was calculated for each drug condition and included as a covariate in sensitivity analyses. Full ASL preprocessing details are provided in the Supplementary Methods.

### 2.7 Behavioural analyses

#### 2.7.1 Primary and sensitivity analyses

Behavioural outcomes included accuracy, false alarm rate (FAR), sensitivity (d’), criterion bias, and latency/reaction time. Signal Detection Theory was used to distinguish sensitivity from response bias: d’ reflects the ability to discriminate target from non-target stimuli, whereas criterion bias reflects the tendency to respond that a target is present [37]. Detailed calculations for each parameter are provided in the Supplementary Methods.

Behavioural analyses were conducted using R Studio Version 2025.05.0+496. Generalised linear mixed-effects models with a binomial distribution and logit link were used for accuracy and FAR, and linear mixed effects models were used for d’, criterion bias, and reaction times. Models included group (Study 1) or drug condition (Study 2), emotion, and their interaction, with participant ID included as a random effect. Statistical significance was set at *p*<0.05, with Bonferroni correction for emotion-specific effects.

Sensitivity analyses additionally examined the effects of age, sex, IQ, and anxiety in Study 1, and anxiety, fatigue and drug order in Study 2. Further model specifications are provided in the Supplementary Methods.

#### 2.7.2 Exploratory misattribution analyses

Directional emotion misattribution errors were compared between CHR-P individuals and HCs in Study 1, and between diazepam and placebo in Study 2 using generalised linear mixed effects models with a binomial distribution and logit link function. Full model specifications are provided in the Supplementary Methods.

### 2.8 fMRI primary and secondary analyses

First-level analyses were conducted in SPM12 [56]. Regressors modelled neutral, threat (anger + fear), and non-threat (happy + sad) expressions [42], with a fixation cross matched to faces on perceptual qualities as the implicit baseline [42, 48, 49]. Contrasts of interest were threat>neutral, non-threat>neutral, neutral>fixation and threat>non-threat. Follow-up threat>fixation and non-threat>fixation contrasts were examined to determine whether group differences in threat>neutral and non-threat>neutral were driven primarily by responses to neutral expressions.

At the second-level, task-related activation was examined using one sample t-tests for the primary contrast of interest, threat>neutral, across groups (Study 1) and in the placebo condition (Study 2). Group differences in Study 1 and diazepam effects in Study 2 were examined using two-sample and paired sample t-tests, respectively, for each task contrast.

Pre-defined bilateral regions-of-interest (ROI) included the amygdala, ACC, fusiform gyrus, hippocampus, insula, and mPFC [59]. ROI analyses used small volume correction (SVC) in SPM12, with significance defined as peak-level family-wise error rate-corrected *p*_FWE_<0.05 within each ROI; false discovery rate (FDR) correction across ROIs is additionally reported.

Supplementary whole-brain analyses assessed peak- and cluster-level FWE corrected *p*_FWE_<0.05 with a voxel-wise threshold of *p*_uncorr_<0.001. Threat-related processing constituted the primary hypotheses. Further modelling, covariate, correction, and effect size details are provided in the Supplementary Methods.

## 3. Results

### 3.1 Study 1

#### 3.1.1 Participant Characteristics

Participant demographic and clinical characteristics are detailed in Table 1. The CHR-P and HC groups differed significantly in ethnicity distribution and IQ, with higher IQ in HCs. CHR-P individuals also had higher positive and negative symptom, depression and anxiety scores, and lower social functioning than HCs.

**Table 1.**
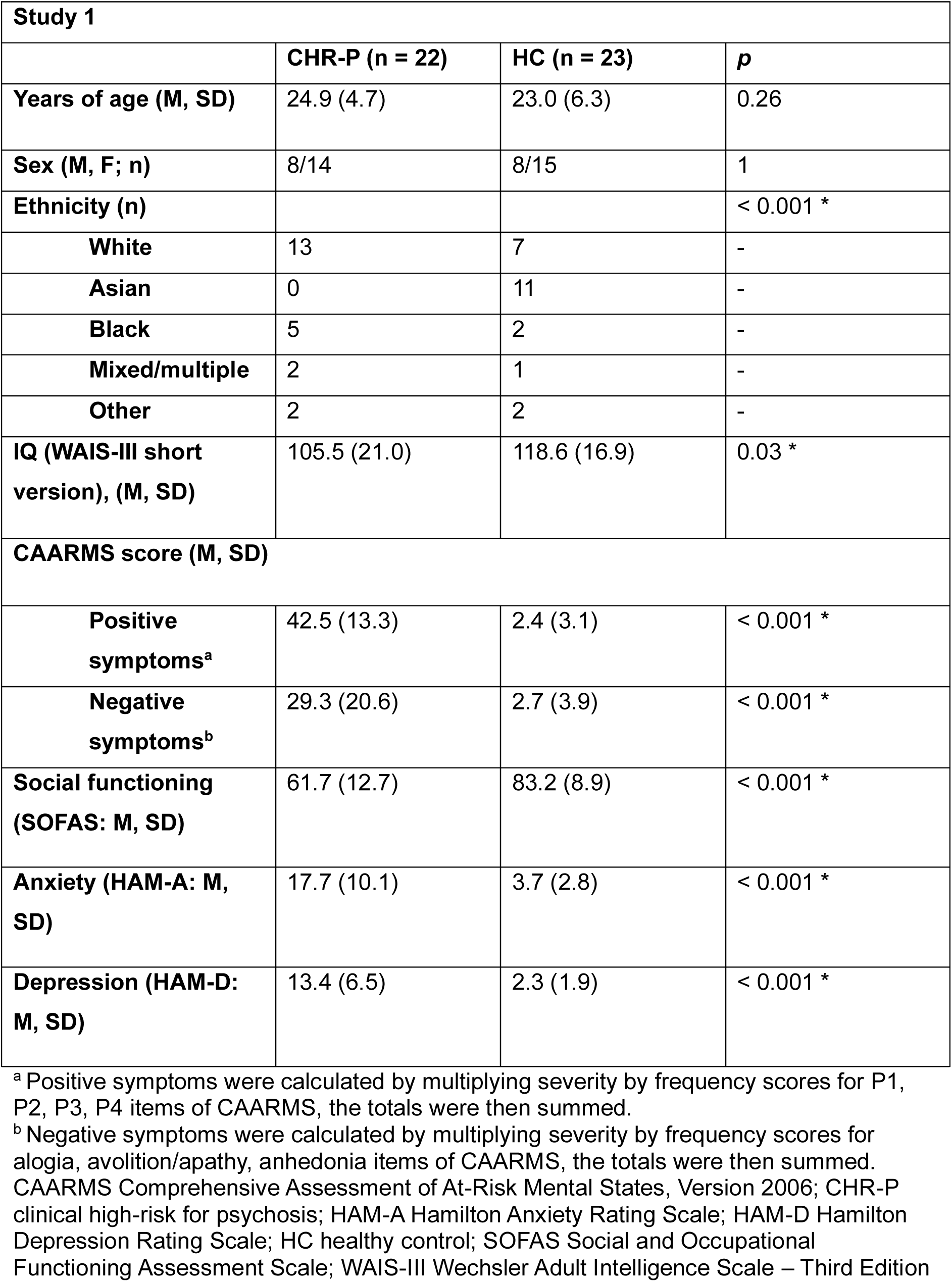
Participant characteristics in Study 1.

#### 3.1.2 Behavioural Results

There were no significant Group or Group x Emotion effects for accuracy, FAR, sensitivity or criterion bias (all *p*’s>0.05, see Supplementary Figures S2A-D and Supplementary Table S2). Individuals at CHR-P had significantly longer reaction times (M=1.97 seconds, 95% CI=[1.85, 2.09]) than HCs (M=1.75 seconds, 95% CI=[1.64, 1.87]) across facial expressions (F(1, 43)=6.88, *p*=0.01, d=-0.78; Figure 1A). This finding was unchanged in sensitivity analyses and after excluding overlapping participants (n=4) to account for repeated task exposure (Supplementary Tables S3-S4).

**Figure 1.**
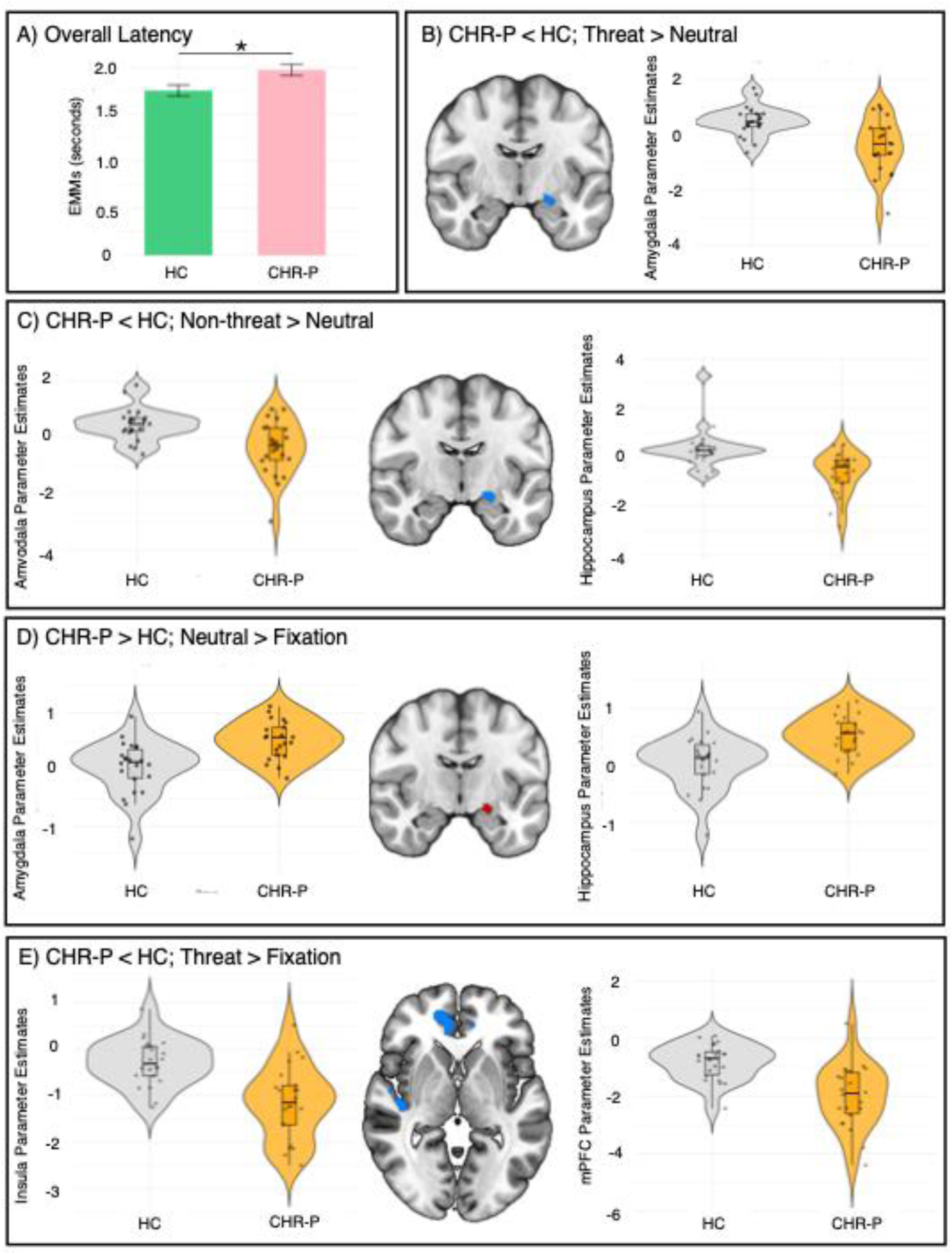
Study 1 Findings: A) Latency estimated marginal means (seconds), error bars represent standard error; B) Lower amygdala activation for threat>neutral in CHR-P relative to HCs; C) Lower amygdala and hippocampal activation for non-threat>neutral in CHR-P relative to HCs; D) Greater amygdala and hippocampal activation for neutral>fixation in CHR-P relative to HCs; E) Lower insula and mPFC activation for threat>fixation in CHR-P relative to HCs; *p<0.05; differences in brain activation were considered significant at pFWE<0.05.

No significant group differences in directional misattributions were observed (Supplementary Table S5).

#### 3.1.3 fMRI Results

##### Task activation

Across groups, threat>neutral elicited significant activation within the a priori corticolimbic network, including in the fusiform gyrus and insula. Supplementary whole-brain analyses are reported in Supplementary Table S6.

##### CHR-P versus HC

Significant peaks were identified near the amygdala-hippocampal border. Compared with HC, CHR-P individuals showed lower amygdala activation for threat>neutral, lower amygdala and hippocampus activation for non-threat>neutral, and greater amygdala and hippocampus activation for neutral>fixation (Table 2, Figure 1B-D). No significant group differences were found in the remaining ROIs or for threat>non-threat.

**Table 2.** Regions of activation differences between individuals at CHR-P and HC in Study 1.

|  | Side | MNI Coordinates (mm) |  |  | K | t | p<sub>FWE</sub> | p<sub>FDR</sub> | d |
|---|---|---|---|---|---|---|---|---|---|
|  |  | x | y | z |  |  |  |  |  |
| **CHR-P<HC** |  |  |  |  |  |  |  |  |  |
| Threat>Neutral |  |  |  |  |  |  |  |  |  |
| Amygdala-hippocampal border<sup>a</sup> | L | -22 | -8 | -12 | 10 | 3.77 | 0.03 | 0.17 | 1.16 |
| Non-threat>Neutral |  |  |  |  |  |  |  |  |  |
| Amygdala-hippocampal border<sup>a</sup> | L | -22 | -8 | -12 | 7 | 4.09 | 0.01 | 0.07 | 1.25 |
| Amygdala-Hippocampal border<sup>b</sup> | L | -20 | -8 | -12 | 32 | 4.16 | 0.04 | 0.11 | 1.27 |
| Threat>Fixation |  |  |  |  |  |  |  |  |  |
| Insula | R | 40 | -18 | 4 | 48 | 4.94 | 0.01 | 0.05 | 1.51 |
| Medial prefrontal cortex | R | 8 | 44 | -4 | 173 | 4.86 | 0.02 | 0.06 | 1.48 |
| **CHR-P>HC** |  |  |  |  |  |  |  |  |  |
| Neutral>Fixation |  |  |  |  |  |  |  |  |  |
| Amygdala-hippocampal border<sup>a</sup> | L | -22 | -8 | -12 | 8 | 4.06 | 0.01 | 0.08 | 1.25 |
| Amygdala-hippocampal border<sup>b</sup> | L | -20 | -10 | -12 | 27 | 4.12 | 0.04 | 0.13 | 1.27 |

<sup>a</sup>Amygdala SVC; <sup>b</sup>Hippocampus SVC
ROI results are reported at peak-level FWE correction within each ROI. Following additional false discovery rate (FDR) correction across ROIs; no ROI findings survived this correction.

To determine whether group differences in the threat/non-threat>neutral contrasts reflected reduced responses to emotional expressions or greater responses to neutral expressions, follow-up analyses examined threat>fixation and non-threat>fixation. CHR-P individuals showed lower insula and mPFC activation for threat>fixation (Table 2, Figure 1E) with no significant group differences for non-threat>fixation. However, ROI findings did not survive additional FDR correction across ROIs.

Supplementary whole-brain and sensitivity analyses are reported in Supplementary Table S7 and the Supplementary Results.

### 3.2 Study 2

#### 3.2.1 Participant Characteristics

Participant demographic and clinical characteristics are detailed in Table 3, a CONSORT flowchart is displayed in Supplementary Figure S14. The sample comprised 23 participants.

**Table 3.** Participant characteristics in Study 2.

| **Study 2** |  |
|---|---|
| **CHR-P (n = 23)** |  |
| **Age (years): M (SD)** | 24.2 (4.8) |
| **Sex n: M/F** | 8/15 |
| **Ethnicity (n)** |  |
| **White** | 12 |
| **Asian** | 2 |
| **Black** | 5 |
| **Mixed/multiple** | 4 |
| **Other** | 0 |
| **IQ (WAIS-III short version): M (SD)** | 98.9 (21.3) |
| **CAARMS: M (SD)** |  |
| **Positive symptoms<sup>a</sup>** | 46.2 (13.2) |
| **Negative symptoms<sup>b</sup>** | 29.5 (25.3) |
| **Social functioning (SOFAS: M, SD)** | 56.2 (9.9) |
| **Anxiety (HAM-A: M, SD)** | 17.0 (8.7) |
| **Depression (HAM-D: M, SD)** | 13.7 (7.1) |
| **Global GM CBF (ml/100g/min): M (SD)** |  |
| **Placebo** | 54.86 (12.96) |
| **Diazepam** | 50.04 (11.41) |

<sup>a</sup> Positive symptoms were calculated by multiplying severity by frequency scores for P1, P2, P3, P4 items of CAARMS, the totals were then summed.
<sup>b</sup> Negative symptoms were calculated by multiplying severity by frequency scores for alogia, avolition/apathy, anhedonia items of CAARMS, the totals were then summed.
CAARMS Comprehensive Assessment of At-Risk Mental States, Version 2006; CHR-P clinical high-risk for psychosis; HAM-A Hamilton Anxiety Rating Scale; HAM-D Hamilton Depression Rating Scale; HC healthy control; SOFAS Social and Occupational Functioning Assessment Scale; WAIS-III Wechsler Adult Intelligence Scale – Third Edition

#### 3.2.2 Behavioural Results

There were no significant Drug or Drug x Emotion effects for accuracy, FAR, sensitivity, criterion bias, or reaction time in the primary analyses (all *p*>0.05; Supplementary Figures S3A-E; Supplementary Table S8).

There was no significant effect of diazepam on d’ (sensitivity) in the full sample (F(1, 106.42)=0.77, *p*=0.38, d_av_=0.13; Figure 2A). In a secondary analysis adjusting for baseline anxiety, which excluded two participants with missing HAM-A data, sensitivity was significantly higher under diazepam (M=3.07, 95% CI=[2.83, 3.31]) than placebo (M=2.90, 95% CI=[2.67, 3.14]) across all expressions (F(1, 95.83)=4.56, *p*=0.04, d_av_=0.29). Repeating the unadjusted analysis in the same reduced sample revealed a similar effect (F(1, 95.96)=4.50, *p=*0.04, d_av_=0.30; Figure 2A; diazepam M=3.07, 95% CI=[2.83, 3.30], placebo M=2.91, 95% CI=[2.67, 3.14]), indicating that this finding was attributable to participant exclusion rather than adjustment for anxiety. Cook’s D test was applied to rule out the influence of potential outliers; this analysis identified one of these participants as potentially influential. In a leave-one-out analysis, only the exclusion of this participant yielded a nominally significant drug effect on sensitivity (F(1, 101)=3.94, *p*=0.0499, d_av_=0.27; Figure 2A; diazepam M=3.05, 95% CI=[2.81, 3.27], placebo M=2.89, 95% CI=[2.67, 3.12]). Sensitivity and influence analyses are reported in Supplementary Tables S9-S11.

**Figure 2.**
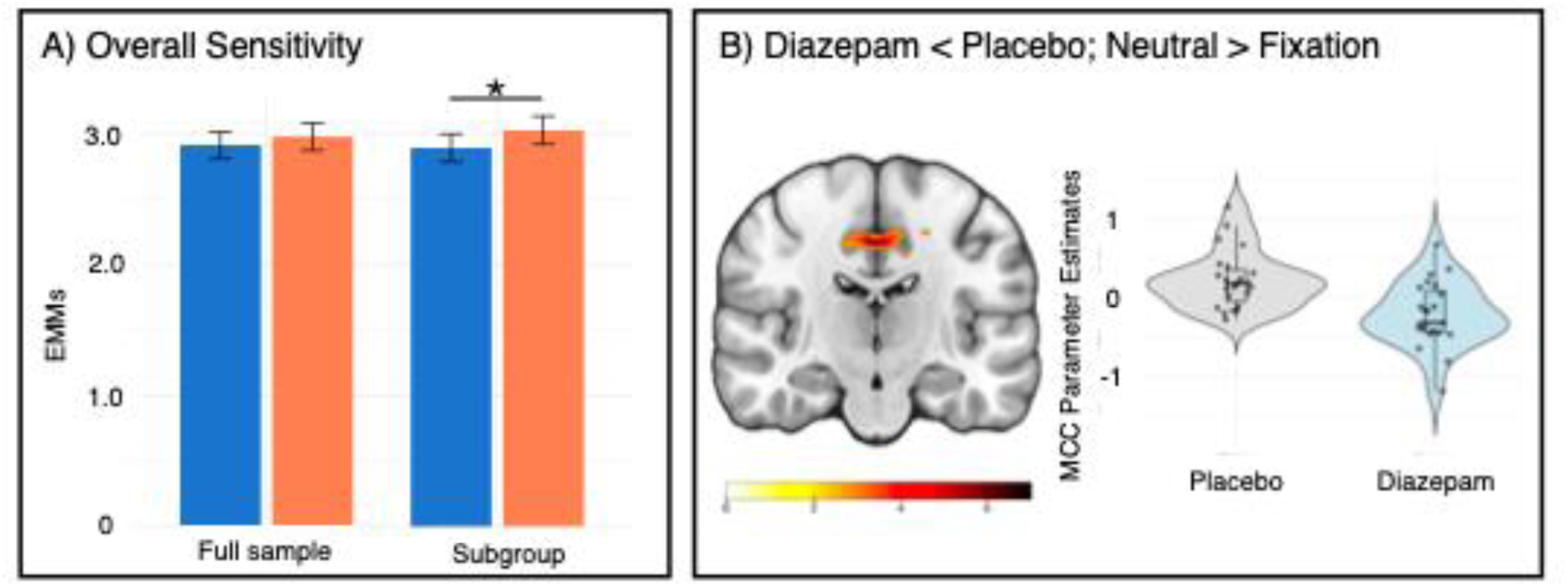
Study 2 Findings: A) Overall sensitivity estimated marginal means for the full unadjusted d’ analysis (n = 23) and the unadjusted d’ subgroup sensitivity analysis (n = 22), error bars represent standard error; B) Attenuated midcingulate cortex activation for neutral>fixation under diazepam relative to placebo; * = p<0.05, differences in brain activation were considered significant at pFWE<0.05.

No significant drug effects on directional misattribution were observed (Supplementary Table S12).

#### 3.2.3 fMRI Results

##### Task Activation

Under placebo, threat>neutral elicited significant activation within the a priori corticolimbic ROIs, including the fusiform gyrus. Supplementary whole-brain results are reported in Supplementary Table S13.

##### Diazepam versus Placebo

No significant drug effects were observed for the threat>neutral, non-threat>neutral, or threat>non-threat contrasts in the pre-specified ROIs, nor for threat>fixation and non-threat>fixation in follow-up analyses. These findings remained non-significant after adjustment for global CBF.

In a secondary whole-brain voxel-wise analysis adjusting for global CBF, diazepam significantly reduced midcingulate cortex activation for neutral>fixation (*k*=343; x=2, y=-18, z=44; t=7.28; peak-level *p*FWE=0.03, cluster-level *p*FWE=0.04, d_av_=-1.12, Figure 2B). No significant drug effects were observed for other contrasts.

## 4. Discussion

Our main findings were that individuals at CHR-P showed slower responses during facial emotion recognition and altered neural responses to emotional and neutral facial expressions. Specifically, CHR-P individuals showed longer reaction times during FER compared to HC, but no significant differences in accuracy, FAR, sensitivity, or criterion bias. At the neural level, CHR-P individuals showed greater activation near the amygdala-hippocampal border to neutral expressions and lower insula and mPFC activation for threat relative to fixation. In Study 2, diazepam produced no significant behavioural effect in the primary analysis and no significant effects within the pre-specified ROIs. Secondary analyses suggested greater overall sensitivity (d’) following diazepam in a reduced sample and attenuated midcingulate cortex (MCC) responses to neutral expressions after adjustment for global CBF. Overall, while diazepam did not normalise the alterations observed in CHR-P, the findings suggest that aspects of emotion processing may be sensitive to GABAergic modulation.

### Case-control findings

Individuals at CHR-P showed slower reaction times across emotions but no significant differences in accuracy, FAR, sensitivity, criterion bias, or directional misclassifications, contrary to our hypothesis of heightened behavioural threat processing. Similar slower reaction times have been reported in SSDs [42, 60, 61] and may reflect greater processing demands despite otherwise comparable FER performance. The absence of other behavioural findings is consistent with previous CHR-P studies that report subtle or absent impairments despite altered neural activation [18, 20, 62, 63]. Indeed, FER deficits may become more pronounced in individuals with greater vulnerability to subsequent illness progression [11], although longitudinal investigations are required.

At the neural level CHR-P individuals showed greater activation near the amygdala-hippocampal border to neutral expressions and reduced activation in these regions for threat>neutral and non-threat>neutral. Similarly heightened amygdala/hippocampal responses to neutral facial expressions have been reported [64–67], and interpreted as reflecting reduced emotional responsivity [13], heightened responsivity to neutral stimuli [32, 68–70], or both [71]. In our follow-up analyses, threat>fixation and non-threat>fixation revealed no corresponding amygdala or hippocampus group differences, suggesting that the reduced activation observed for threat>neutral and non-threat>neutral was primarily driven by greater responses to neutral faces. Elevated responses to neutral facial expressions may reflect aberrant assignment of emotional salience [21] although neutral expressions remain relatively understudied in CHR-P because they are commonly used as a baseline condition [17].

Separate post-hoc analyses showed lower insula and mPFC activation for threat>fixation. Both regions have established roles in threat and aversive processing [72, 73], although previous findings primarily implicate the anterior insula. Similar reductions have been reported in SSDs [74], raising the possibility that altered threat-related recruitment may be present at the CHR-P stage.

### Experimental Medicine Study

Contrary to our hypotheses, diazepam had no significant effects on emotion processing accuracy, FAR, sensitivity, criterion bias, or latency in the primary analysis. In a secondary analysis adjusting for baseline anxiety, sensitivity was higher under diazepam, but this analysis excluded two participants with missing HAM-A data. Repeating the unadjusted analysis in this reduced sample yielded a similar effect, suggesting that finding was attributable to participant exclusion rather than anxiety adjustment. Post-hoc influence analyses identified one of these participants as potentially influential, and exclusion of this participant alone also resulted in a nominally significant effect. Thus, these findings suggest a potential increase in overall emotion processing sensitivity under diazepam, although this effect was sensitive to participant inclusion and should be interpreted with caution.

No diazepam effects were observed for criterion bias, directional misattribution, or emotion-specific outcomes. If replicated, the selective effect on sensitivity could suggest improved discrimination rather than altered response tendencies, although the processes underlying this behavioural finding remain inconclusive. Previous work in healthy individuals similarly reported that 5 mg diazepam did not affect emotion processing accuracy or latency [75], whereas higher doses of diazepam have altered recognition of threatening emotions but also produced sedation [35, 36]. The 5 mg dose used here may have limited detectable behavioural effects while reducing sedation-related confounding.

At the neural level, diazepam did not significantly affect threat processing within the pre-specified ROIs. However, in a secondary whole-brain analysis adjusting for global CBF, diazepam reduced MCC activation to neutral expressions. The MCC has been implicated in salience processing [76], and aberrant salience attribution to irrelevant or ambiguous stimuli is proposed to play an important role in psychosis risk [77–79]. This finding raises the possibility that GABAergic modulation influences neural responses to socially ambiguous stimuli in CHR-P. Previous studies have also reported altered MCC activation during emotion recognition in CHR-P, although not specifically to neutral expressions [18, 80]. Although the regional effect differed from those in Study 1, both findings suggest that neural responses to neutral facial expressions warrant further investigation in CHR-P.

The absence of ROI effects differs from previous pharmacological imaging studies in healthy individuals using 10 mg diazepam [34] and first-degree relatives of individuals with schizophrenia using 1mg alprazolam [49], which reported benzodiazepine-related modulation of amygdala, hippocampus, ACC and insula responses. Differences may reflect variation in benzodiazepine dose or type, sample characteristics, task design, or fMRI contrast specification. For example, the alprazolam study [49] collapsed emotional and neutral expressions into a single overall condition, increasing the number of trials contributing to the contrast of interest. Higher benzodiazepine doses may also produce greater neural effects but increase sedation-related confounding. Overall, the absence of effects within the pre-specified corticolimbic ROIs provides little evidence that 5 mg diazepam robustly modulated the hypothesised circuitry. The whole-brain MCC finding nevertheless suggests that neural responses to neutral expressions may be sensitive to acute GABAergic modulation. Across both studies, neutral facial expressions emerged as a potentially informative condition for understanding altered social-emotional processing in CHR-P.

To our knowledge, this was the first study to apply Signal Detection Theory to characterise FER in individuals at CHR-P and the first to investigate the acute effects of diazepam on FER and its neural correlates in this population. The use of SDT across both studies enabled emotion processing performance to be characterised beyond conventional accuracy metrics, by distinguishing sensitivity from criterion bias. Nevertheless, several limitations should also be considered. Some significant findings arose from post-hoc, sensitivity, or supplementary analyses. In Study 1, threat>fixation and non-threat>fixation analyses were conducted post-hoc; in Study 2, the behavioural effect emerged only in reduced-sample analyses, and the MCC effect only after adjustment for global CBF in a secondary analysis. These findings should therefore be interpreted with caution and require replication. The ER40 also showed ceiling or floor effects for some accuracy and FAR metrics, which may have limited sensitivity to subtle behavioural differences and constrained derived d’ and criterion bias estimates. Combining anger and fear into a threat category, and sadness and happiness into a non-threat category, increased trial numbers but may have obscured emotion-specific effects. Both studies included relatively small sample sizes, 13 CHR-P participants contributed to both studies, there were significant differences in ethnicities between CHR-P and HCs in Study 1, and Study 2 lacked a HC group, preventing assessment of whether diazepam shifted responses towards a normative pattern. Finally, the 5 mg diazepam dose was selected to minimise sedation; future pharmaco-fMRI studies could use dose-response designs and more selective GABA_A_ receptor compounds to better delineate GABAergic effects on emotion processing in CHR-P.

## Conclusion

Emotion processing in individuals at CHR-P is associated with altered corticolimbic activation, particularly elevated amygdala-hippocampal responses to neutral facial expressions, despite largely comparable behavioural performance to healthy controls. Diazepam did not significantly affect behavioural performance in the primary analysis or activation within the pre-specified corticolimbic ROIs, although secondary analyses suggested increased overall emotion sensitivity and reduced MCC responses to neutral expressions. These findings highlight the potential relevance of neutral, socially ambiguous facial expressions in CHR-P and suggest that aspects of emotion processing may be sensitive to GABAergic modulation.

## Data Availability

Case-control and diazepam-placebo whole-brain fMRI contrast data will be made available upon publication. Any other data and associated code are available from the corresponding author on reasonable request.

## ACKNOWLEDGEMENTS

We would like to thank the participants who took part in these studies. We would also like to thank Beate Haege (South London and Maudsley NHS Foundation Trust), OASIS clinical team members, Eugenii A. Rabiner for help with study set-up at Perceptive.Inc, and the radiographers at the Centre for Neuroimaging Sciences.

## AUTHOR CONTRIBUTIONS

NCJ conducted the behavioural and neuroimaging analyses and prepared the figures, with methodological input from PCTH and OOD. NCJ wrote the original draft of the manuscript. PBL, AK, NRL and SRK contributed to participant recruitment and data acquisition. JD contributed to neuroimaging data acquisition, while NV and JD supported participant recruitment. AdM, TJS and PFP provided clinical oversight. GM, EC and MAM supervised the analyses, interpretation of findings and manuscript development. GM conceived and designed the studies, secured funding and oversaw the research programme. All authors critically revised the manuscript for important intellectual content, approved the final version for publication, and agree to be accountable for all aspects of the work.

## DECLARATION OF GENERATIVE AI

During the preparation of this work, the author(s) used ChatGPT for code generation and proofreading and editing. The author(s) reviewed and edited the output as needed and take full responsibility for the content of the published article.

