## Supplementary Material for "Facial Emotion Processing and GABAergic Modulation in Individuals at Clinical High-Risk for Psychosis"

Supplementary Information

### Supplementary Methods: Study 1

**Visit Procedures** (see table below)

Participants attended a screening visit and a single PET-MR scanning session. In the screening visit, demographic information, basic medical history, and clinical and cognitive assessments were collected. The scanning visit described formed part of a wider PET-MR protocol involving bolus administration of [^11^C]Ro15-4513 with arterial blood sampling, and 70 minutes dynamic acquisition. Following completion of PET acquisition, participants underwent fMRI while completing the Penn Emotion Recognition Task (ER40) [1-3]; only ER40 fMRI data are analysed in the present study. The radiotracer dose of [^11^C]Ro15-4513 was not expected to have any pharmacological effects on ER40 fMRI.

*Clinical assessments:*

- CHR-P psychopathology assessed via the Comprehensive Assessment of At-Risk Mental States (CAARMS) [4]
- Anxiety and depression assessed via Hamilton Anxiety and Depression Rating Scales (HAM-A; HAM-D) [5, 6]
- Social functioning, disability, adjustment, and interactions, emotional empathy assessed via Social and Occupational Functioning Scale (SOFAS) [7], Global Functioning Social and Role (GF-S/R) [8], World Health Organisation Disability Assessment Schedule (WHODAS-II) [9], Social Adjustment Scale-Self Report (SAS-SR) [10], Interpersonal Sensitivity Measure (IPSM) [11], Emotional Perspective Taking Task (EPTT) [12]
- Childhood maltreatment assessed via the Childhood Trauma Questionnaire (CTQ) [13]
- Impacts of stressors on life events assessed via Holmes-Rahe Life Stress Inventory [14]
- Perceptions of recent stress assessed via Perceived Stress Scale (PSS) [15]
- Dispositional mindfulness assessed via the Kentucky Inventory of Mindfulness Skills (KIMS) [16]

*Cognitive assessments:*

- IQ assessed via the Wechsler Adult Intelligence Scale (WAIS-III) short version [17]
- Premorbid IQ assessed via the National Adult Reading Test [18]
- Cognitive flexibility and visuospatial attention assessed via the Trail-making test A and B [19]

*Subjective states pre- and post-scan:*

- Anxiety and other subjective states assessed via Visual Analogue Mood Scale (VAMS)
- Physical manifestations of anxiety assessed via the Bodily Symptoms Scale (BSS) [20]

*fMRI tasks:*

- Penn Emotion Recognition Task (ER40)
  - The ER40 is a computerised cognitive task to assess static facial affect recognition [1-3]. In an extension of ER40, sixty colour photographs are shown in a pseudo-randomised event-related design displaying faces expressing one of four basic emotions (anger, fear, happiness, sadness) or neutral expressions for 4000ms. Variable intervals ranging from 2000-12000ms, weighted towards shorter intervals, displaying a complex fixation cross matched on facial perceptual qualities were interspersed throughout the trials. Face stimuli were balanced for sex, age and ethnicity. Participants were required to identify the displayed emotion by button-press. In statistical analyses, conditions of interest included threat (anger + fear), non-threat (happy + sad), and neutral [1].
- Fractal n-back task

| **-100 min** | Arrive at the centre |
| --- | --- |
| **-90 min** | ID, Consent and pre-scan checklist. During waiting subject given pre-scan self-report measures to complete. Basic physical examination (weight, height, blood pressure, and standard systems exam). |
| **-70 min** | HCG urine pregnancy test; drug screen |
| **-60min** | Arterial and venous cannula inserted. Self-report measures to complete. |
| **-10 min** | Trip to toilet |
| **T = 0** | Imaging commences, with PET acquisition commencing at T=0 and lasting 70 minutes; total imaging duration is up to 130 minutes. |
|  | A tracer dose of 450 MBq (^11^C)Ro15-4513 is given |
| **+ 130 min** | End scan. Arterial line is removed and aftercare is given. Subject escorted to recovery room. Subject provided with snack and refreshment. Change into own clothes. |
| **45 min** | Post-scan self-report measures |

*Clinical characteristics scores*

- Positive symptoms were calculated by multiplying severity x frequency scores for P1, P2, P3, P4 CAARMS positive subscales, these were then summed
- Negative symptoms were calculated by multiplying severity x frequency scores for alogia, avolition/apathy, anhedonia CAARMS negative subscales, these were then summed

*MRI acquisition*

MRI data were acquired at Perceptive Discovery, London, using a 3T GE SignaTM PET-MR General Electric scanner. T1-weighted structural images were acquired using an Inversion Recovery Spoiled Gradient Echo (IR-SPGR) sequence (TR/TE/TI = 6.992/2.996/400 ms; flip angle = 11°; matrix = 256x256, FoV = 256; in-plane resolution = 1.0x1.0 mm; slice thickness = 1 mm; 200 slices). Functional images were acquired using a gradient-echo pulse sequence (TR/TE = 2000/30 ms; flip angle = 80°; matrix = 64x64; FoV = 192; in-plane resolution = 3.0x3.0 mm; slice thickness = 3.6 mm; 35 slices; 210 volumes).

*ER40 fMRI*

For both studies, quality control of raw images was conducted using MRIQC [21]. Motion and slice timing correction, co-registration, and normalisation to the MNI template were conducted using fMRIPrep 23.1 [22], and visually inspected using CheckReg function in SPM12 [23]. Further quality control of individual maps was conducted using an in-house script [24]: for participants with >10% of volumes with framewise displacement (FD) > 1mm, correlations between the FD time series and each task condition regressor were examined to identify task-correlated head motion. In the case of a significant correlation, participants were excluded from second-level analyses of contrasts containing that condition. Participants with >20% of volumes with FD > 1mm were excluded from all imaging analyses. Scrubbing was conducted using an in-house script [24], with volumes of FD > 1 mm modelled as regressors of no interest in the first-level GLM. Individual functional images were spatially smoothed with a 6mm FWHM Gaussian kernel using SPM12 [23]. To ensure that motion scrubbing did not disproportionately affect individual emotions within each condition (threat, non-threat), we analysed the overlap between motion flagged volumes (FD > 1 mm) and stimulus presentations for anger and fear, and happiness and sadness. Across this, the difference in number of presentations without overlap with flagged volumes was no more than three trials within each condition, suggesting broadly comparable representation of the emotions. No participants were excluded entirely, though contrast-specific exclusions were made and are detailed in Supplementary Table 1.

*Behavioural analyses*

Behavioural outcomes included accuracy, false alarm rate (FAR), sensitivity (d’), and criterion bias, as per SDT [25], and latency/reaction time. SDT is a framework for analysing decision-making in the presence of uncertainty [25], an approach particularly relevant to psychosis, where FER abnormalities have been associated with reduced sensitivity and altered criterion biases, including biases towards attributing threat-related emotions to neutral expressions [26-29]. The calculation of d’ and criterion bias were based on Macmillan and Creelman (1990) [25]; the calculation of adjusted Hit Rates and FARs were based on Corwin (1994) [30] to negate for infinite z-scores. d’ reflects the ability to discriminate between two stimuli (i.e. threat from neutral, threat from non-threat, non-threat from neutral), with a high d’ value indicating a good ability to discriminate, and a low value indicating a poor ability to discriminate [25]. Criterion bias is a measure of bias, reflecting the amount of evidence required before a response is provided for a target stimulus; no bias has a value of 0, negative values indicate a greater tendency to judge a target as present, even when limited evidence is available, whereas positive values indicate that a participant requires stronger evidence before responding that the target is present [25].

Behavioural analyses were conducted using R Studio Version 2025.05.0+496. General linear mixed effects models with binomial error distributions and logit links were used to analyse accuracy and FAR were used to assess the effects of group, emotion, and their interaction on accuracy and FARs. Group, emotion, and group x emotion were included as fixed effects; subject ID was included as a random effect to account for repeated measures. Accuracy and FARs were separately modelled with binomial error distributions and logit link function. Separate linear mixed effects models were employed to assess the effect of group, emotion, and their interaction on d’ and criterion bias. Group, emotion, and group x emotion were included as fixed effects, with subject ID as a random effect to account for repeated measures.

We conducted sensitivity analyses for completeness. Sex, age, IQ and anxiety scores measured via the Hamilton Anxiety Rating Scale (HAM-A) [5] were included as covariates. Statistical significance was set at *p* = 0.05, with p-values for emotion-specific effects corrected using the Bonferroni method.

*Exploratory misattribution analyses*

To explore the directionality of emotional categorisation errors, we conducted a misattribution analysis. For each participant, confusion matrices were constructed to quantify the frequency with which each target emotion (threat, non-threat), or neutral expression was assigned to each response emotion or neutral expression. Misattributions were compared between CHR-P and HCs. Analyses were conducted using generalised linear mixed effects models with a binomial error distribution logit link function. Group, direction of misattribution, and group x direction were modelled as fixed effects, subject ID was included as a random effect. Statistical significance was set at *p* = 0.05, with p-values for direction-specific effects corrected using the Bonferroni method.

*fMRI analyses*

Imaging data were analysed using SPM12 [23]. First-level analyses were performed on each subject using GLM. The BOLD signal was modelled using a first-level GLM. The task regressors of interest were neutral, threat (anger + fear), non-threat (happy + sad) [3]. These regressors were convolved with a canonical haemodynamic response function to estimate parameter estimates per subject. A fixation cross matched to faces on perceptual qualities served as the implicit baseline [1-3]. Six head motion parameters (three rotations and three translations) and binary regressors corresponding to scrubbed volumes were included as regressors of no interest. Low frequency drift was removed using a high-pass filter with a 128-second cut-off. First level contrasts were calculated as threat > neutral, non-threat > neutral, neutral > fixation, threat > non-threat; follow-up analyses were conducted using contrasts of threat > fixation, non-threat > fixation to determine whether observed group differences in threat > neutral and non-threat > neutral were primarily driven by responses to neutral expressions.

Second-level analyses were also conducted in SPM12. One sample t-tests were performed for the primary contrast of interest, threat > neutral, to confirm task-related activation across groups. Two-sample t-tests were used to examine group differences for each task contrast. To account for participant characteristic differences, analyses were repeated including IQ as a covariate.

Pre-defined bilateral anatomical masks were generated for key FER-related regions-of-interest, including the amygdala, anterior cingulate cortex (ACC), fusiform gyrus, hippocampus, insula, and medial prefrontal cortex (mPFC) [31], using the Automated Anatomical Labelling (AAL) atlas from the WFU_Pickatlas toolbox in SPM12 (Supplementary Figure S1). Region of interest (ROI) analyses were conducted using small volume correction (SVC) in SPM12, with statistical significance defined as peak-level family-wise error rate-corrected *p* (*p*_FWE_ ) < 0.05 within each ROI, with additional false discovery rate (FDR) correction across ROIs reported with a significance threshold of *p_FDR_* _<_ 0.05.

Whole-brain analyses were conducted for completeness; significance was set at *p*_FWE_ < 0.05 at the cluster-level, with a height threshold of *p*_uncorr_ < 0.001. Threat-related processing constituted the primary hypotheses; no additional correction was applied across contrasts. Cohen’s d was calculated to quantify the magnitude of between-subject effects.

*Automated Anatomical Atlas ROI Mask Generation*

- Anterior cingulate cortex = Cingulum_Ant_L + Cingulum_Ant_R
- Bilateral amygdala = Amygdala_L + Amygdala_R
- Fusiform gyrus = Fusiform_L + Fusiform_R
- Hippocampus = Hippocampus_L + Hippocampus_R
- Insula = Insula_L + Insula_R
- Medial prefrontal cortex = Frontal_Sup_Medial_L + Frontal_Sup_Medial_R

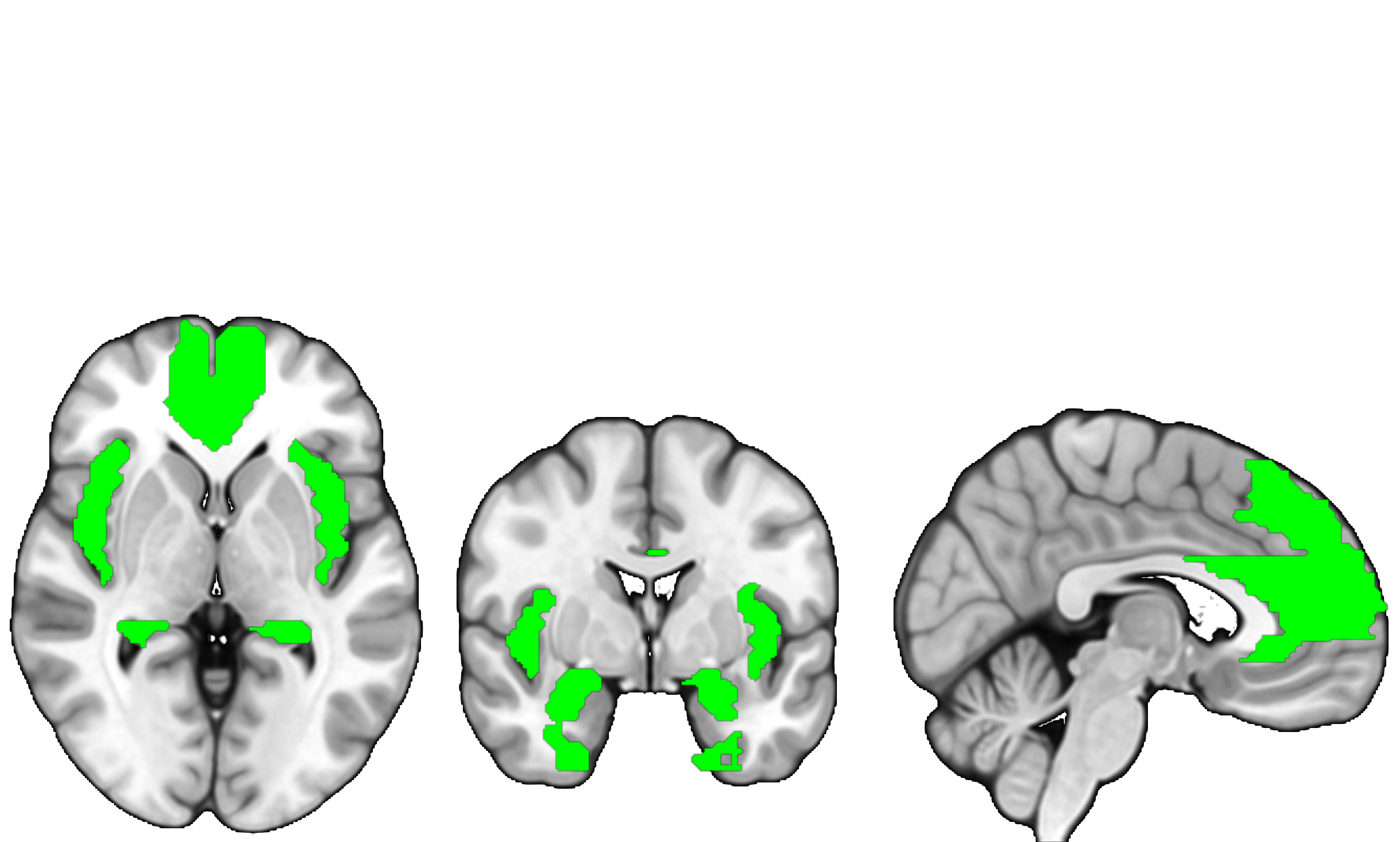

**Figure S1**: Corticolimbic ROI mask.

### Supplementary Methods: Study 2

Study 2: A power calculation was conducted for the study’s main outcome measure using G*Power on MRS glutamate and GABA levels collected in our centre [32]. The calculation indicated that 24 subjects per group were required to detect a significant change with a power of 0.8 (alpha = 0.05, two tailed).

Participants underwent a screening visit and two MRI sessions, once under a single oral dose of 5 mg diazepam and once under oral placebo (50 mg ascorbic acid), with a minimum 3-week washout period between scanning visits. At the screening visit, demographic information, basic medical history, and clinical and cognitive assessments were obtained. At the scanning visits, participants were administered either diazepam or placebo in a randomised and counterbalanced order 60 minutes prior to MRI scanning, for scanning to commence at diazepam’s t_max_ [33], and completed the ER40 while undergoing blood oxygen-level dependent (BOLD) fMRI. Study 2 was not informed by patient or public involvement.

Participants and all study staff (other than outpatient hospital pharmacy staff) were blinded. Diazepam and placebo pills were re-packaged into opaque gel capsules before collection by the study team. A computerised randomisation list was generated by an external researcher to control for order effects; treatment order was concealed from all study staff (other than outpatient hospital pharmacy staff). Randomisation was implemented using a Williams design [34]; participants were allocated to either AB or BA treatment orders, where AB = diazepam followed by placebo and BA = placebo followed by diazepam.

**Visit Procedures** (see table below)

Recruitment took place between 5^th^ September 2019 and 24^th^ March 2023, with the last outcomes collected on 24^th^ March 2023.

*Clinical assessments:*

- CHR-P psychopathology assessed via CAARMS
- Anxiety and depression assessed via HAM-A and HAM-D
- Social functioning, disability, adjustment, and interactions, emotional empathy assessed via SOFAS, GF-S/R, WHODAS-II, SAS-SR, IPSM, EPTT
- Childhood maltreatment assessed via CTQ
- Dispositional mindfulness assessed via KIMS
- Concomitant medication use was recorded

*Cognitive assessments:*

- IQ assessed via the WAIS-III short version
- Premorbid IQ assessed via NART
- Trail-making test A and B

*Subjective states pre- and post-scan:*

- Anxiety and other subjective states assessed via Visual Analogue Mood Scale (VAMS)
- Physical manifestations of anxiety assessed via the Bodily Symptoms Scale (BSS)

*fMRI tasks:*

- ER40
- Fractal n-back task

|  | Screening Visit  (~2 hours total) | Visit 1 (~3.5 hours total) | Visit 2 (~3.5 hours total) |
| --- | --- | --- | --- |
| Participant information and informed consent | X |  |  |
| Sociodemographics | X |  |  |
| Inclusion/exclusion criteria, including MRI safety questionnaire | X |  |  |
| Blood sampling | X |  |  |
| Clinical/cognitive assessment | X |  |  |
| Subjective measures |  | X | X |
| Physical examination |  | X | X |
| Benzodiazepine / Placebo administration |  | X | X |
| MRI Scanning |  | X | X |
| Adverse event monitoring |  | X | X (plus additional follow-up call after 7 days) |

*Clinical characteristics scores*

- Positive symptoms were calculated by multiplying severity x frequency scores for P1, P2, P3, P4 CAARMS positive subscales, these were then summed
- Negative symptoms were calculated by multiplying severity x frequency scores for alogia, avolition/apathy, anhedonia CAARMS negative subscales, these were then summed

*Adverse event monitoring*

Safety reporting followed NRES guidelines (<http://www.hra.nhs.uk/resources/during-and-after-your-study/progress-and-safety-reporting/>). Adverse events were monitored by designated study clinicians. No serious adverse events were reported.

*MRI acquisition*

MRI data were acquired at the Centre for Neuroimaging Sciences, King’s College London, using a General Electric MR750 3.0 T MR scanner with a 32-channel head coil. T1-weighted structural images were acquired using an IR-SPGR sequence (TR/TE/TI = 7.31/3.02/400 ms; flip angle = 11° matrix = 256x256, FoV = 270; in-plane resolution = 1.05x1.05 mm; slice thickness = 1.2mm; 196 slices). Functional images were acquired using a single-shot gradient-echo EPI sequence (TR/TE = 2000/30 ms; flip angle = 75°; matrix = 64x64; FoV = 211; in-plane resolution = 3.30x3.30 mm; slice thickness = 3; 41 slices, 251 volumes).

Regional CBF was measured using a 3D pseudo-continuous arterial spin labelling sequence with a multi-shot 3D Fast Spin Echo Stack of Spirals as used in previous CHR-P studies from our group [35-37]. Full details are described in our previous publication [38] (TR/TE = 4,923/11.088 ms; post-labelling delay = 2025 ms; FoV = 240; reconstructed resolution = 2x2x3 mm; 56 slices).

*Stopping guidelines*

End of study was defined as the last participant’s final scanning visit.

*Behavioural analyses*

Behavioural analyses were conducted using R Studio Version 2025.05.0+496. General linear mixed effects models with a binomial error distribution and logit link were used to analyse accuracy and FAR were used to assess the effects of drug, emotion, and their interaction on accuracy and FARs. Drug, emotion, and drug x emotion were included as fixed effects; subject ID was included as a random effect to account for repeated measures. Accuracy and FARs were separately modelled with binomial error distributions and logit link function. Separate linear mixed effects models were employed to assess the effect of drug, emotion, and their interaction on d’ and criterion bias. Drug, emotion, and drug x emotion were included as fixed effects, with subject ID as a random effect to account for repeated measures.

We conducted sensitivity analyses for completeness. Anxiety scores measured via HAM-A [5], fatigue measured via the Bodily Symptoms Scale [20], and drug/placebo order effects were included as covariates. Statistical significance was set at *p* = 0.05, with p-values for emotion-specific effects corrected using the Bonferroni method.

Post-hoc influential diagnostics were conducted using Cook’s distance with a threshold of 4/n [39] to identify influential participants. Leave-one-subject out-analyses were performed across the full sample to assess the robustness of model effects.

*Exploratory misattribution analyses*

Misattributions were compared between drug and placebo. Analyses were conducted using generalised linear mixed effects models with a binomial error distribution and logit link function. Drug, direction of misattribution, and drug x direction were modelled as fixed effects, subject ID was included as a random effect. Statistical significance was set at *p* = 0.05, with p-values for direction-specific effects corrected using the Bonferroni method.

*fMRI analyses*

Imaging data were analysed using SPM12 [23]. First-level analyses were performed on each subject using GLM, The BOLD signal was modelled using a first-level GLM. The task regressors of interest were neutral, threat (anger + fear), non-threat (happy + sad) [3]. These regressors were convolved with a canonical haemodynamic response function to estimate parameter estimates per condition. A fixation cross matched to faces on perceptual qualities served as the implicit baseline [1-3]. Six head motion parameters (three rotations and three translations) and binary regressors corresponding to scrubbed volumes were included as regressors of no interest. Low frequency drift was removed using a high-pass filter with a 128-second cut-off. First level contrasts were calculated as threat > neutral, non-threat > neutral, neutral > fixation, threat > non-threat; follow-up analyses were conducted using contrasts of threat > fixation, non-threat > fixation for completeness.

Second-level analyses were also conducted in SPM12. One sample t-tests were performed for the primary contrast of interest, threat > neutral, to confirm task-related activation in the placebo condition. Paired sample t-tests were used to examine diazepam effects for each task contrast. ROI analyses were conducted using SVC in SPM12, with statistical significance defined as peak-level family-wise error rate-corrected *p* (*p*_FWE_ ) < 0.05 within each ROI, with additional FDR correction across ROIs reported with a significance threshold of *p_FDR_* _<_ 0.05.

To account for potential effects of task order and of diazepam-related sedation, analyses were repeated including task order and participant-reported fatigue, as reported via the Bodily Symptoms Scale [20], as covariates. In these same CHR-P participants we previously demonstrated that 5 mg diazepam significantly reduced global CBF [38]. Given that drug-induced changes in global CBF may influence estimates of task-related BOLD activation, Study 2 analyses were repeated including global CBF as a covariate (per drug condition).

Whole-brain analyses were conducted for completeness; significance was set at *p*_FWE_ < 0.05 at the cluster-level, with a height threshold of *p*_uncorr_ < 0.001. Threat-related processing constituted the primary hypotheses; no additional correction was applied across contrasts. Cohen’s d (d_av_) was calculated as the mean within-participant difference between diazepam and placebo, divided by the pooled standard deviation of the drug/placebo conditions.

### Study 1 and Study 2 Participant Exclusions

Table S1: Study 1 and Study 2 participant exclusions

| Study | Analysis | Group/Condition | Initial n | Excluded n | Included n | Exclusion reasons |
| --- | --- | --- | --- | --- | --- | --- |
| Study 1 | Behavioural | HC | 24 | 1 | 23 | Missing data |
|  |  | CHR-P | 24 | 2 | 22 | Task disengagement; missing data |
|  | fMRI: Threat > Neutral | HC | 24 | 2 | 22 | Neutral excessive motion; missing data |
|  |  | CHR-P | 24 | 3 | 21 | Neutral excessive motion, Threat excessive motion; Missing data |
|  | fMRI: Non-threat > Neutral | HC | 24 | 2 | 22 | Neutral excessive motion; Missing data |
|  |  | CHR-P | 24 | 2 | 22 | Neutral excessive motion; Missing data |
|  | fMRI: Neutral > Fixation | HC | 24 | 3 | 21 | Neutral excessive motion, Fixation excessive motion; Missing data |
|  |  | CHR-P | 24 | 2 | 22 | Neutral excessive motion; Missing data |
|  | fMRI: Threat > Non-threat | HC | 24 | 1 | 23 | Missing data |
|  |  | CHR-P | 24 | 2 | 22 | Threat excessive motion; Missing data |
|  | fMRI: Threat > Fixation | HC | 24 | 2 | 22 | Fixation excessive motion; Missing data |
|  |  | CHR-P | 24 | 2 | 22 | Threat excessive motion; Missing data |
|  | fMRI: Non-threat > Fixation | HC | 24 | 2 | 22 | Fixation excessive motion; Missing data |
|  |  | CHR-P | 24 | 1 | 23 | Missing data |
| Study 2 | Behavioural | Placebo | 24 | 1 | 23 | Missing data |
|  |  | Diazepam | 24 | 3 | 21 | Missing data |
|  | fMRI: Threat > Neutral | Placebo | 24 | 1 | 23 | Missing data |
|  |  | Diazepam | 24 | 1 | 23 | Missing data |
|  | fMRI: Non-threat > Neutral | Placebo | 24 | 1 | 23 | Missing data |
|  |  | Diazepam | 24 | 1 | 23 | Missing data |
|  | fMRI: Neutral > Fixation | Placebo | 24 | 1 | 23 | Missing data |
|  |  | Diazepam | 24 | 1 | 23 | Missing data |
|  | fMRI: Threat > Non-threat | Placebo | 24 | 1 | 23 | Missing data |
|  |  | Diazepam | 24 | 1 | 23 | Missing data |
|  | fMRI: Threat > Fixation | Placebo | 24 | 1 | 23 | Missing data |
|  |  | Diazepam | 24 | 1 | 23 | Missing data |
|  | fMRI: Non-threat > Fixation | Placebo | 24 | 1 | 23 | Missing data |
|  |  | Diazepam | 24 | 1 | 23 | Missing data |

### Supplementary Results: Study 1

#### Statistical Results: Study 1

Table S2: Study 1: Effects of Group and Group x Emotion on Accuracy, False Alarm Rate, Sensitivity, and Criterion bias

| Outcome | Group, statistic (*p*) | Group x Emotion, statistic (*p*) |
| --- | --- | --- |
| Accuracy | *χ²* = 0.7683 (0.38) | *χ²* = 0.9655 (0.62) |
| False Alarm Rate | *χ²* = 0.0422 (0.84) | *χ²* = 3.1968 (0.20) |
| Sensitivity | F = 0.1473 (0.70) | F = 0.2184 (0.80) |
| Criterion Bias | F = 0.0131 (0.91) | F = 0.7552 (0.47) |

Table S3:Study 1 Sensitivity Analyses: Effects of Group and Group x Emotion

| Outcome | Covariate | Group, statistic (*p*) | Group x Emotion, statistic (*p*) |
| --- | --- | --- | --- |
| Accuracy | Anxiety | *χ²* = 1.8936 (0.17) | *χ²* = 1.5502 (0.46) |
|  | Age | *χ²* = 0.5171 (0.47) | *χ²* = 0.9616 (0.62) |
|  | Sex | *χ²* = 0.8283 (0.36) | *χ²* = 0.9771 (0.61) |
|  | IQ | *χ²* = 0.1867 (0.67) | *χ²* = 1.0169 (0.60) |
| False Alarm Rate | Anxiety | *χ²* = 0.0850 (0.77) | *χ²* = 3.1736 (0.20) |
|  | Age | *χ²* = 0.1807 (0.67) | *χ²* = 3.1860 (0.20) |
|  | Sex | *χ²* = 0.0378 (0.85) | *χ²* = 3.1946 (0.20) |
|  | IQ | *χ²* = 0.7307 (0.39) | *χ²* = 4.4737 (0.11) |
| Sensitivity | Anxiety | F = 0.5844 (0.45) | F = 0.3037 (0.74) |
|  | Age | F = 0.0493 (0.83) | F = 0.2184 (0.80) |
|  | Sex | F = 0.1486 (0.70) | F = 0.2184 (0.80) |
|  | IQ | F = 0.0297 (0.86) | F = 0.1542 (0.86) |
| Criterion Bias | Anxiety | F = 0.1405 (0.71) | F = 1.0617 (0.35) |
|  | Age | F = 0.0029 (0.96) | F = 0.7496 (0.47) |
|  | Sex | F = 0.0130 (0.91) | F = 0.7506 (0.47) |
|  | IQ | F = 0.0057 (0.94) | F = 0.9222 (0.40) |
| Latency | Anxiety | F = 2.4110 (0.13) | F = 0.8860 (0.42) |
|  | Age | F = 7.7371 (0.008)* | F = 1.0249 (0.36) |
|  | Sex | F = 8.5772 (0.006)* | F = 1.0249 (0.36) |
|  | IQ | F = 4.0801 (0.05)* | F = 1.0986 (0.34) |

Table S4: Study 1 SDT outcomes, excluding for n = 4 repeat participants

| Outcome | Group, statistic (*p*) | Group x Emotion, statistic (*p*) |
| --- | --- | --- |
| Accuracy | *χ²* = 1.4753 (0.22) | *χ²* = 2.3406 (0.31) |
| False Alarm Rate | *χ²* = 0.0901 (0.76) | *χ²* = 1.9738 (0.37) |
| Sensitivity | F = 0.1332 (0.72) | F = 0.5594 (0.57) |
| Criterion Bias | F = 0.0313 (0.86) | F = 0.6606 (0.51) |
| Latency | F = 11.0454 (0.002)* | F = 1.4739 (0.24) |

Table S5: Study 1: Effects of Group and Group x Direction on Directional Misattributions

| Covariate | Group, statistic (*p*) | Group x Direction, statistic (*p*) |
| --- | --- | --- |
| Unadjusted | F = 0.0381 (0.85) | F = 0.5956 (0.70) |
| Anxiety | F = 0.2148 (0.65) | F = 0.6496 (0.66) |
| Age | F = 0.0052 (0.94) | F = 0.5956 (0.70) |
| Sex | F = 0.0298 (0.86) | F = 0.5956 (0.70) |
| IQ | F = 0.0426 (0.84) | F = 0.7094 (0.62) |

#### Behavioural Figures: Study 1

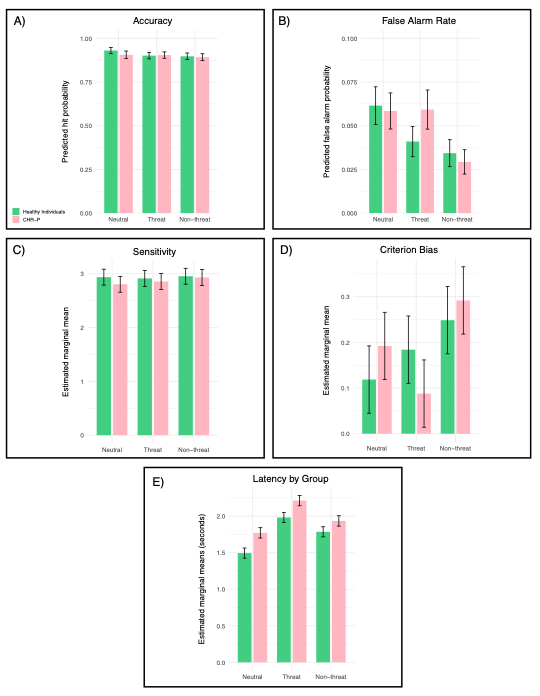

Figure S2: Study 1 behavioural results: A) Accuracy predicted probabilities; B) False alarm rate predicted probability; C) Sensitivity estimated marginal means; D) Criterion bias estimated marginal means; E) Latency estimated marginal means. Error bars represent standard error.

#### fMRI Task Activation: Study 1

Table S6: Study 1: Task-related brain activation for threat > neutral across all participants

|  | Side | MNI Coordinates | | | *cluster-extent* | *t* | *pFWE* |
| --- | --- | --- | --- | --- | --- | --- | --- |
|  |  | x | y | z |  |  |  |
| **Whole-brain** | | | | | | | |
| **Activation (CHR-P + HC)** | | | | | | | |
| Primary visual cortex | R | 14 | -90 | 4 | 2945 | 9.64 | <0.001 |
| Supplementary motor cortex | L | -4 | 4 | 58 | 3717 | 6.95 | <0.001 |
| Superior parietal lobule | L | -24 | -60 | 60 | 1097 | 6.04 | <0.001 |
| Inferior occipital gyrus | L | -26 | -94 | 0 | 323 | 6.03 | 0.022 |
| Precentral gyrus | R | 30 | -6 | 52 | 548 | 6 | 0.001 |
| Inferior frontal gyrus | L | -36 | 20 | 8 | 513 | 5.78 | 0.001 |
| Superior parietal lobule | R | 24 | -62 | 60 | 333 | 5.55 | 0.018 |
| **Deactivation (CHR-P + HC)** | | | | | | | |
| Whole-brain | | | | | | | |
| Angular gyrus | R | 54 | -66 | 40 | 1300 | 6.72 | <0.001 |
| Angular gyrus | L | -54 | -60 | 46 | 1316 | 6.37 | <0.001 |
| Posterior cingulate cortex | R | 16 | -44 | 34 | 1339 | 5.60 | <0.001 |
| Superior frontal gyrus | L | -16 | 40 | 34 | 537 | 5.24 | 0.001 |
| Superior frontal gyrus | R | 16 | 40 | 56 | 465 | 4.65 | 0.003 |
| **ROI** | | | | | | | |
| **Activation (CHR + HC)** | | | | | | | |
| Fusiform gyrus | R | 26 | -80 | -8 | 309 | 6.61 | <0.001 |
| Insula | L | -32 | 26 | -2 | 327 | 5.75 | 0.001 |
| Insula | R | 36 | 24 | -2 | 238 | 4.84 | 0.013 |

#### fMRI analysis: Study 1

Supplementary whole-brain voxel-wise analysis of threat>fixation revealed a significant cluster of hypoactivation in the lingual gyrus (CWM; *k* = 1,697; x = -12, y = -76, z = -6; t = 4.86; cluster-level *p*FWE<0.001) in the CHR-P group. There were no other significant differences in activation between groups.

#### fMRI Sensitivity analysis: Study 1

Table S7: Regions of activation differences between individuals at CHR-P and HCs, adjusted for IQ (Study 1)

|  | Side | MNI Coordinates (mm) | | | K | t | p_FWE_ | d |
| --- | --- | --- | --- | --- | --- | --- | --- | --- |
|  |  | x | y | z |  |  |  |  |
| **CHR-P < HC** | | | | | | | | |
| Threat > Neutral | | | | | | | | |
| Amygdala/hippocampal border | L | -22 | -8 | -12 | 18 | 4.05 | 0.02 | 1.25 |
| Non-threat > Neutral | | | | | | | | |
| Amygdala/hippocampal border | L | -22 | -8 | -12 | 4 | 3.75 | 0.03 | 1.14 |
| Threat > Fixation | | | | | | | |  |
| Medial prefrontal cortex^a^ | R | 10 | 44 | -4 | 185 | 4.60 | 0.04 | 1.40 |
| Lingual gyrus | L | -22 | -58 | -4 | 375 | 3.70 | 0.01 (cluster), 0.64 (peak) | 1.13 |
| Superior frontal gyrus medial segment^a^ | R | 10 | 44 | -4 | 293 | 4.60 | 0.047 (cluster), 0.58 (peak) | 1.40 |
| Anterior cingulate cortex | L | -4 | 32 | 14 | 63 | 4.44 | 0.03 | 1.35 |
| **CHR-P > HC** | | | | | | | | |
| Neutral > Fixation | | | | | | | | |
| Amygdala/hippocampal border | L | -22 | -8 | -12 | 9 | 4.52 | 0.004 | 1.39 |
| Amygdala/hippocampal border | L | -20 | -10 | -12 | 53 | 4.45 | 0.02 | 1.37 |

^a^Medial prefrontal cortex ROI and superior frontal gyrus medial segment whole-brain findings share the same peak coordinate

### Supplementary Results: Study 2

#### Statistical Results: Study 2

Table S8: Study 2: Effects of Drug and Drug x Emotion on Accuracy, False Alarm Rate, Sensitivity, Criterion bias, and Latency

| Outcome | Drug, statistic (*p*) | Drug x Emotion, statistic (*p*) |
| --- | --- | --- |
| Accuracy | *χ²* = 0.1056 (0.75) | *χ²* = 3.6686 (0.16) |
| False Alarm Rate | *χ²* = 0.8206 (0.37) | *χ²* = 4.5466 (0.10) |
| Sensitivity | F = 0.7670 (0.38) | F = 0.0132 (0.99) |
| Criterion Bias | F = 0.0642 (0.80) | F = 2.0553 (0.13) |
| Latency | F = 0.8494 (0.36) | F = 0.0753 (0.93) |

Table S9: Study 2 Sensitivity Analyses: Effects of Drug and Drug x Emotion, *p < 0.05

| Outcome | Covariate | Drug, statistic (*p*) | Drug x Emotion, statistic (*p*) |
| --- | --- | --- | --- |
| Accuracy | Anxiety | *χ²* = 0.1261 (0.72) | *χ²* = 3.5984 (0.17) |
|  | Order | *χ²* = 0.1057 (0.75) | *χ²* = 3.6688 (0.16) |
|  | Fatigue | *χ²* = 0.1503 (0.70) | *χ²* = 3.6547 (0.16) |
| False Alarm Rate | Anxiety | *χ²* = 0.8686 (0.35) | *χ²* = 4.4597 (0.11) |
|  | Order | *χ²* = 0.8211 (0.36) | *χ²* = 4.5467 (0.10) |
|  | Fatigue | *χ²* = 0.9355 (0.33) | *χ²* = 4.5400 (0.10) |
| Sensitivity | Anxiety | F = 4.5591 (0.04)* | F = 0.1706 (0.84) |
|  | Order | F = 0.7693 (0.38) | F = 0.0132 (0.99) |
|  | Fatigue | F = 0.8377 (0.36) | F = 0.0131 (0.99) |
| Criterion Bias | Anxiety | F = 0.0067 (0.93) | F = 1.6560 (0.20) |
|  | Order | F = 0.0638 (0.80) | F = 2.0392 (0.13) |
|  | Fatigue | F = 0.0632 (0.80) | F = 2.0390 (0.13) |
| Latency | Anxiety | F = 0.5336 (0.47) | F = 0.0724 (0.93) |
|  | Order | F = 0.8736 (0.35) | F = 0.2451 (0.78) |
|  | Fatigue | F = 0.8815 (0.35) | F = 0.2449 (0.78) |

Table S10: Study 2 SDT outcomes, excluding for n = 9 repeated participants

| Outcome | Drug, statistic (*p*) | Drug x Emotion, statistic (*p*) |
| --- | --- | --- |
| Accuracy | *χ²* = 0.0657 (0.80) | *χ²* = 1.9045 (0.39) |
| False Alarm Rate | *χ²* = 0.4519 (0.50) | *χ²* = 2.1624 (0.34) |
| Sensitivity | F = 0.3688 (0.55) | F = 0.1183 (0.89) |
| Criterion Bias | F = 0.0613 (0.81) | F = 0.9775 (0.38) |
| Latency | F = 0.8719 (0.35) | F = 0.0096 (0.99) |

Table S11: Influential participant effects on leave one out subject analyses, as identified by Cook's d

| Outcome | SubjectID | Cook’s distance | Relative to threshold* | LOSO drug *p-*value | LOSO drug*emotion *p-*value |
| --- | --- | --- | --- | --- | --- |
| False alarm rate | SUBJECT01 | 0.392320820 | 2.25584472 | 0.278 | 0.102 |
| False alarm rate | SUBJECT02 | 0.284908364 | 1.63822310 | 0.531 | 0.140 |
| False alarm rate | SUBJECT03 | 0.250103447 | 1.43809482 | 0.108 | 0.236 |
| Accuracy | SUBJECT04 | 0.271771852 | 1.56268815 | 0.960 | 0.0663 |
| Accuracy | SUBJECT05 | 0.180784094 | 1.03950854 | 0.867 | 0.229 |
| Sensitivity | SUBJECT03 | 0.226986939 | 1.30517490 | 0.04993* | 0.90371 |
| **Threshold = 0. 173913* [39]*; LOSO = leave-one-subject-out analysis. N.B. SubjectIDs have been anonymised* | | | | | |

Table S12: Study 2: Effects of Drug and Drug x Direction on Directional Misattributions

| Covariate | Drug, statistic (*p*) | Drug x Direction, statistic (*p*) |
| --- | --- | --- |
| Overall model | F = 0.0386 (0.84) | F = 0.4226 (0.83) |
| Anxiety | F = 0.4446 (0.51) | F = 0.4186 (0.84) |
| Order | F = 0.0342 (0.85) | F = 0.4214 (0.83) |
| Fatigue | F = 0.0157 (0.90) | F = 0.4227 (0.83) |

#### Behavioural Figures: Study 2

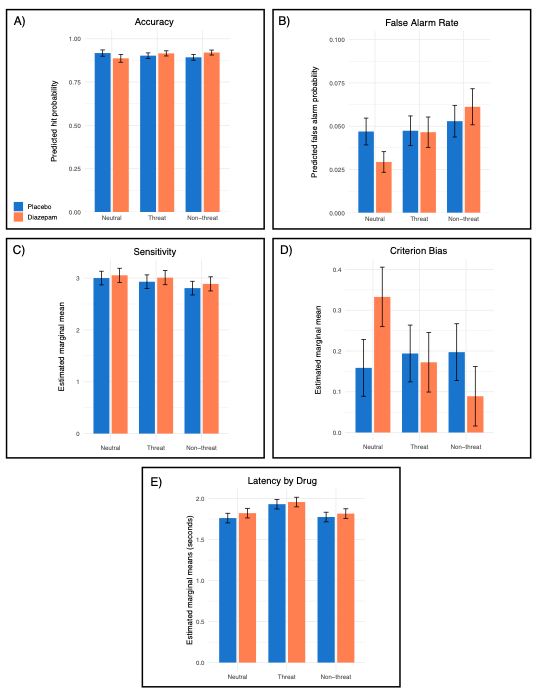

Figure S3: Study 2 behavioural results. A) Accuracy predicted probabilities; B) False alarm rate predicted probability; C) Sensitivity estimated marginal means; D) Criterion bias estimated marginal means; E) Latency estimated marginal means. Error bars represent standard error.

#### fMRI Task Activation: Study 2

Table S13: Study 2: Task-related brain activation for threat > neutral in the placebo condition

|  | Side | MNI Coordinates (mm) | | | *cluster-extent* | *t* | *pFWE* |
| --- | --- | --- | --- | --- | --- | --- | --- |
|  |  | x | y | z |  |  |  |
| **Whole-brain** | | | | | | | |
| **Activation (CHR-P, placebo)** | | | | | | | |
| Superior occipital gyrus | R | 30 | -88 | 20 | 4234 | 8.63 | <0.001 |
| Middle occipital gyrus | L | -28 | -90 | 6 | 1239 | 6.35 | <0.001 |
| Middle frontal gyrus | L | -46 | 22 | 26 | 1875 | 5.32 | <0.001 |
| **Deactivation (CHR-P, placebo)** | | | | | | | |
| Postcentral gyrus | L | -50 | -18 | 46 | 9227 | 8.18 | <0.001 |
| Middle frontal gyrus | R | 32 | 30 | 52 | 468 | 5.65 | 0.03 |
| **ROI** | | | | | | | |
| **Activation (CHR-P, placebo)** | | | | | | | |
| Fusiform gyrus | R | 24 | -78 | -10 | 586 | 7.25 | <0.001 |

### CONSORT Flowchart: Study 2

Table S14: CONSORT Flowchart

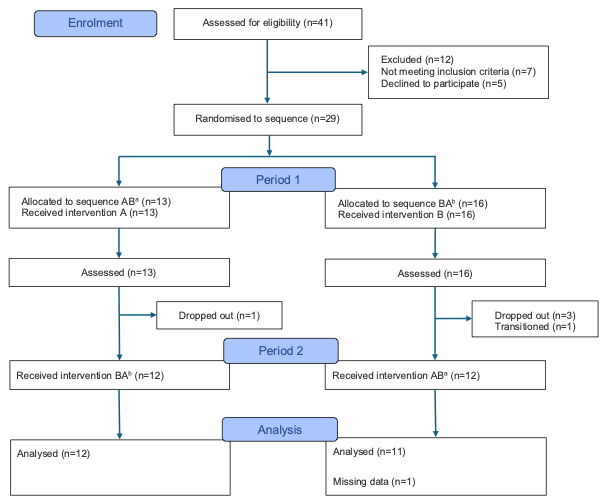

*CONSORT = Consolidated Standards of Reporting Trials* [40, 41].

*^a^Diazepam followed by placebo; ^b^placebo followed by diazepam; all interventions and comparators were administered as intended*
